# Music reshapes basal ganglia neural state dynamics toward a medication-like regime in Parkinson’s disease

**DOI:** 10.64898/2026.08.17.26360495

**Authors:** Sandeep Sathyanandan Nair, Veronika Filyushkina, Kamal R. Chémali, Aratrik Guha, Anna Gamaleya, Alexey Tomskiy, Alexey Sedov, Aasef G. Shaikh

**Author notes:** Corresponding Author: Aasef G. Shaikh,MD,PhD 11100 Euclid Avenue, Cleveland OH 44022 USA.

## Abstract

Parkinson’s disease (PD) is characterized by excessive neural synchronization in the 13–30 Hz beta band within basal ganglia circuits. Conventional therapies, including dopaminergic medication and deep brain stimulation (DBS), reduce beta synchrony while enhancing lower-frequency theta activity. As an adjunct to these treatments, music and auditory rhythms improve motor function in PD, but the neural mechanisms remain unclear. Here, we recorded local field potentials in the subthalamic nucleus (STN) of patients with PD in the medication-off state to test how structured musical elements shape subcortical synchrony. Spectral and neural state-space analyses showed that rhythmic and harmonic components of music produced effects comparable to dopaminergic therapy, suppressing beta-band oscillations while enhancing theta-band activity. These effects were strong in the dorsal sensorimotor STN, whereas the ventral limbic STN showed minor modulation. Directional connectivity analysis further revealed that music-induced beta and theta changes were accompanied by increased cortex-to-STN drive, consistent with top-down recruitment of the cortical-subthalamic hyperdirect pathway. Notably, harmonic consonance produced network-level modulation comparable to, and in some cases greater than, rhythmic entrainment, extending beyond the established framework of beat-based basal ganglia engagement. Together, these findings identify frequency-selective, region-specific, and stimulus-locked mechanisms by which music reshapes pathological basal ganglia activity in PD, providing direct electrophysiological evidence that auditory stimulation can transiently shift subcortical dynamics toward a medication-like state.

## Introduction

Music has long been recognized as more than a cultural expression — it is a complex biological stimulus that engages widespread neural systems spanning cortical, subcortical, and limbic regions simultaneously (Zatorre et al., 2007; Chen et al., 2022; Zaatar et al., 2024). Modern neuroscience conceptualizes music perception as a distributed phenomenon involving perceptual, cognitive, motor, and emotional processing in parallel, with engagement of circuits involved in temporal prediction, reward, and sensorimotor integration (Zatorre & Salimpoor, 2013; Salimpoor et al., 2011).

Because music engages multiple functional domains, it has emerged as a promising therapeutic modality across neurological conditions, including Parkinson’s disease (PD), stroke, and Alzheimer’s disease (Edwards et al., 2023; Thaut & Hoemberg, 2015).

Music-based interventions are thought to strengthen neural pathways involved in sensory processing, motor coordination, emotional regulation, and memory — domains commonly affected by neurodegenerative disorders. In PD specifically, rhythmic auditory stimulation (RAS) has been shown to improve gait velocity, stride length, and cadence, and to reduce freezing of gait episodes (Thaut et al., 1996; Thaut & Hoemberg, 2015; Lan et al., 2026).

PD is characterized by pathological synchronization of beta-band oscillations (13–30 Hz) within the cortico-basal ganglia-thalamic circuit (Hammond et al., 2007; Little & Brown, 2014). This pathological beta synchrony is strongly linked to motor impairment and is attenuated by both dopaminergic medication and deep brain stimulation (DBS) - the two primary therapeutic interventions in PD (Little & Brown, 2014; Kühn et al., 2006; Nair et al., 2022; Nair & Chakravarthy, 2024). Concurrently, dopaminergic therapy is associated with enhancement of lower-frequency activity, particularly in the theta range (4–8 Hz), which has been linked to cognitive control, attentional engagement, and movement initiation (Cavanagh & Frank, 2014; Oswal et al., 2013).

Given music’s capacity to engage distributed neural systems involved in timing, movement, and prediction, its relevance to PD has become increasingly evident. Notably, many of the neural structures most affected in PD including the basal ganglia, supplementary motor area (SMA), and cerebellar circuits are involved in rhythm perception and sensorimotor synchronization during musical engagement (Grahn & Brett, 2007; Bonetti et al., 2024; Chen et al., 2022). This anatomical and functional overlap provides a mechanistic basis for the therapeutic use of music in PD rehabilitation (Edwards et al., 2023; Lan et al., 2026). Within this context, understanding how music interacts specifically with basal ganglia circuitry becomes central to explaining its therapeutic potential in PD.

Music is a multidimensional stimulus composed of distinct structural elements that engage partially dissociable neural systems. Rhythmic structure preferentially recruits motor and basal ganglia circuits involved in temporal prediction and entrainment (Grahn & Brett, 2007; Large & Snyder, 2009). Melodic structure engages auditory cortical networks involved in pitch processing and tonal prediction (Zatorre et al., 2002).

Harmonic consonance interacts with limbic and reward pathways, including dopaminergic midbrain and ventral striatal systems (Blood & Zatorre, 2001; Salimpoor et al., 2011). This raises the possibility that different musical components may differentially modulate basal ganglia dynamics, and that the therapeutic potential of music may not be limited to its rhythmic properties.

Despite growing clinical interest, the neural mechanisms by which music modulates subcortical dynamics in PD remain incompletely characterized. Most prior work has relied on scalp electroencephalography (EEG) or functional MRI, which provide limited spatial resolution within the basal ganglia. We investigated how structured musical stimulation modulates subcortical neural dynamics in PD using post-operative LFP recordings from the externalized STN and GPi leads. We tested the hypothesis that music particularly its rhythmic and harmonic components can induce a shift in basal ganglia activity toward a “medication-like∝ state characterized by suppression of pathological beta oscillations and enhancement of lower-frequency activity. To address this, we combined spectral, time-resolved, state-space, and directionality analyses to characterize the impact of music on neural dynamics across multiple levels of organization, and to identify the cortical sources that may drive these subcortical changes within the basal ganglia.

## Methods

### Participants and recordings

All study procedures were conducted at the Burdenko Neurosurgical Institute, in accordance with applicable institutional and international ethical standards for research involving human participants. The study protocol was reviewed and approved by the Institutional Ethics Committee of the Burdenko Neurosurgical Institute. Before enrollment, all participants received a detailed explanation of the study’s purpose, procedures, potential risks and benefits, and their right to withdraw at any time without affecting their medical care. Written informed consent was obtained from each participant before any study-related procedure. Post-operative LFPs were recorded from 48 sites in the STN in six PD patients after recent DBS implantation. Based on stereotactic targeting and post-operative lead reconstruction in patient-specific MR space, sites were further classified as dorsal STN (sensorimotor; nine sites in six patients), ventral STN (associative/limbic; nine sites in six patients).

### Experimental design

Recordings were obtained during wakefulness in three phases: (1) spontaneous baseline OFF medication, (2) structured music exposure OFF medication, and (3) spontaneous recording ON medication. The music stimulus comprised seven sequential conditions designed to vary tempo, melody, and harmonic valence: Beat1 (60 BPM), Beat2 (120 BPM), Melody, Consonant1 (52 BPM), Consonant2 (120 BPM), Dissonant1 (52 BPM), and Dissonant2 (110 BPM), with 10 s silent intervals between segments. Total trial duration was 174 s, and patient-specific onset times defined analysis windows.

### Data acquisition and preprocessing

LFPs were sampled at 2000 Hz. Signals were visually inspected, and noisy or electrically contaminated segments were excluded. No additional detrending was applied unless stated. Analyses used custom scripts in Python 3.9, MATLAB 2025a, and FieldTrip where appropriate.

### Spectral analysis

Power spectral density (PSD) was estimated with Welch’s method (2 s Hann window, 50% overlap, 2000 Hz sampling). To separate oscillatory from aperiodic activity, PSDs were parameterized with FOOOF/specparam (Donoghue et al., 2020) over 1–45 Hz using a fixed aperiodic mode, peak width limits of 1–12 Hz, up to 8 peaks, minimum peak height 0.1, and threshold 2 SD above the aperiodic fit. The aperiodic component was subtracted from log-PSD to yield flattened spectra; model fits were generally high (R² > 0.90). Band power was then computed in theta (4–8 Hz), alpha (8–13 Hz), beta (13–30 Hz), and gamma (30–45 Hz). Condition effects were expressed as within-subject ΔPSD (dB) relative to each patient’s spontaneous OFF-medication baseline, reducing inter-individual variability due to electrode placement, impedance, and physiology.

### Time–frequency and state-space analysis

Time–frequency representations used short-time Fourier transforms (1024-sample window, 80% overlap). Band-limited power was averaged within canonical bands, normalized to the pre-music baseline, and expressed in dB. To capture joint oscillatory dynamics, each time point was mapped into a 2D state space defined by baseline-relative theta and beta power: Δ*θ*(*t*) = *θ*(*t*) − *θ_baseline_*, Δ*β*(*t*) = *β*(*t*) − *β_baseline_* Condition-specific point clouds were visualized with kernel density estimation and 95% confidence ellipses. The therapeutic state—simultaneous theta enhancement and beta suppression—occupied the upper-left quadrant (Δθ > 0, Δβ < 0). Condition similarity to medication was assessed from centroid distances normalized by pooled covariance, and displacement magnitude from baseline was quantified as

(D(*t*) = √(Δ*θ*^2^(*t*) + Δβ^2^(*t*)).

### Directionality analysis

Directed EEG–LFP connectivity was quantified with the Phase Slope Index (PSI; Nolte et al., 2008), a non-parametric metric robust to volume conduction. Positive PSI indicated cortex leading STN/GPi, and negative PSI indicated subcortex leading cortex. PSI was computed for seven EEG channels (C3, C4, CZ, F3, F4, P3, P4) paired with each LFP contact across canonical frequency bands using FieldTrip (Oostenveld et al., 2011). Music-related change was defined as ΔPSI = *PSI_Music_* − *PSI_Baseline_*.

### Statistical analysis

Non-parametric statistics were used throughout because of small sample size and non-normality. A hierarchical four-test framework was applied. H1/H2 used one-sided Wilcoxon signed-rank tests versus baseline: beta tested for suppression, while theta, alpha, and gamma tested for enhancement, using aperiodic-corrected ΔPSD. H3 used two-sided paired Wilcoxon tests to compare music-induced and medication-induced changes; non-significance indicated statistically comparable effects, although in dorsal STN medication produced greater beta suppression than all music conditions. H4 used a Friedman omnibus test to assess condition structure across patients. Within each group × band, H1/H2 and H3 p-values were corrected separately with Benjamini– Hochberg FDR (Benjamini & Hochberg, 1995). Effect sizes were reported as rank-biserial correlation r (−1 to +1).

## Results

Fig. 1 shows a representative single-patient spectral response to music and dopaminergic medication. In dorsal STN (Fig. 1B1–B2), compared across silence, music, and medication; medication-ON (i.e., therapeutic on medications) caused marked beta suppression (13–30 Hz) in both raw and aperiodic-corrected spectra. Music also modulated beta activity, but less strongly than medication. Theta activity (4– 8 Hz) increased consistently with medication, with little or only modest change during music. Together, these results suggest music affects dorsal STN beta activity, but more weakly than dopaminergic medication.

**Figure 1.**
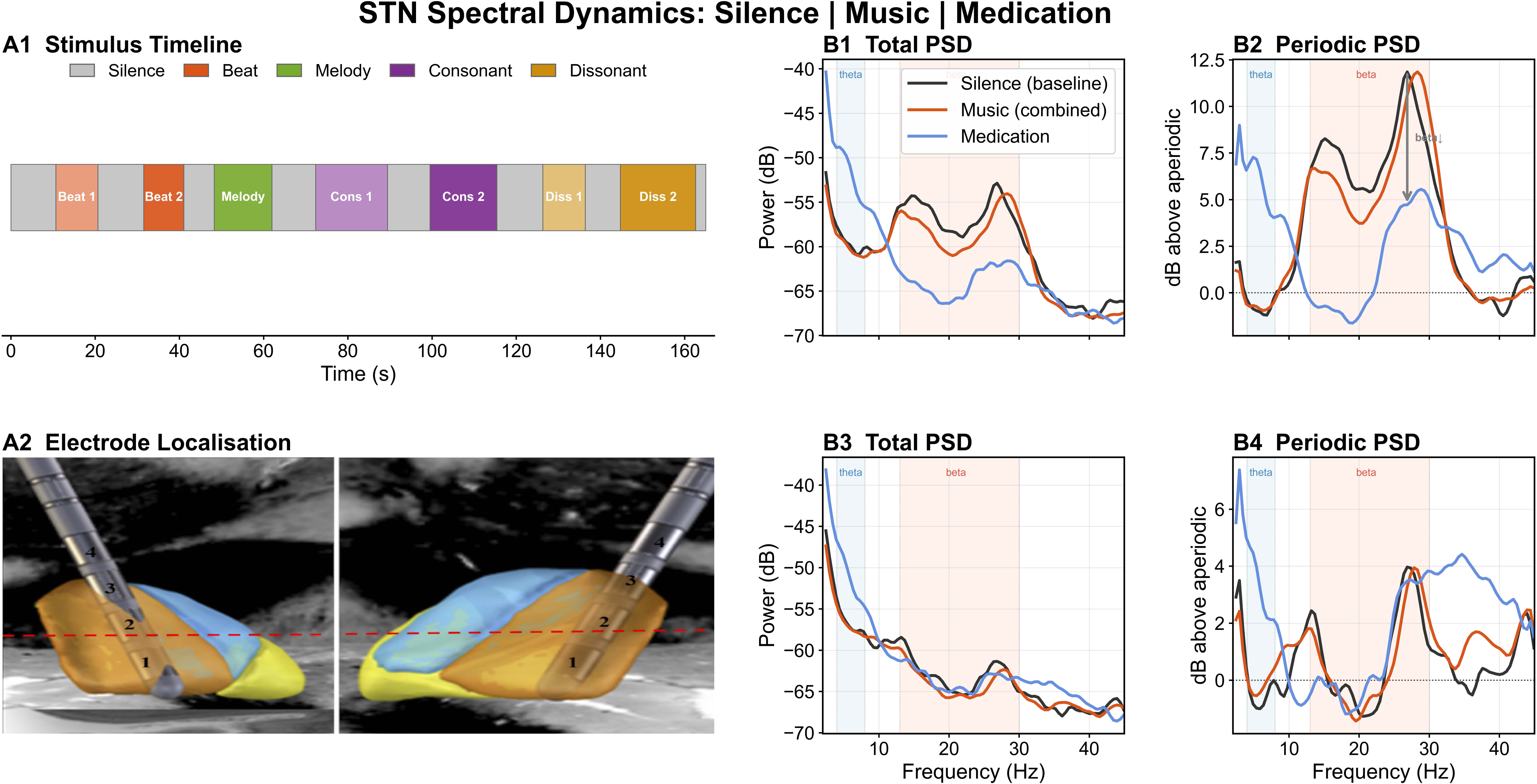
Single-patient illustration of music-induced spectral modulation in subthalamic nucleus (STN) recordings. (A1) Experimental timeline showing the sequence of conditions: baseline (OFF medication), music exposure (beat, melody, consonant), and medication-ON state. (A2) Patient-specific electrode localization in patient-specific MRI indicating dorsal (motor) and ventral (associative/limbic) STN contacts. (B1–B2) Spectral responses from a dorsal STN contact. (B1) Raw power spectral density (PSD) and (B2) aperiodic-corrected (FOOOF-flattened) spectra are shown for the Silence, Music, and Medication conditions. The medication-ON condition produced a clear suppression of beta-band power (13–20 Hz) relative to baseline (arrows). Music conditions also produced measurable spectral deviations from baseline, although the magnitude of modulation was smaller than that observed during medication, suggesting that music exposure can influence STN activity. In contrast, the ventral STN contact (B3-B4) showed no consistent modulation of beta- or theta-band activity across music conditions in either raw or aperiodic-corrected spectra, suggesting regional specificity of the observed effect.

In ventral STN contacts (Fig. 1B3–B4), beta and theta showed no clear or consistent modulation across music conditions. Spectral profiles stayed similar to baseline in both raw and aperiodic-corrected spectra, indicating that effects were specific to dorsal (motor) STN. This pattern—dorsal STN beta suppression during rhythmic and harmonic stimulation and medication, but no ventral effects motivated the group-level analyses.

### Group-level spectral changes across recording sites

Raw PSD cannot be compared directly across patients because electrode placement, contact impedance, and physiology differ. We therefore measured condition effects as aperiodic-corrected PSD change from each patient’s pre-music spontaneous baseline (ΔPSD, dB), where zero (dashed line) indicates no change, negative values suppression, and positive values enhancement (Fig 2).

**Figure 2.**
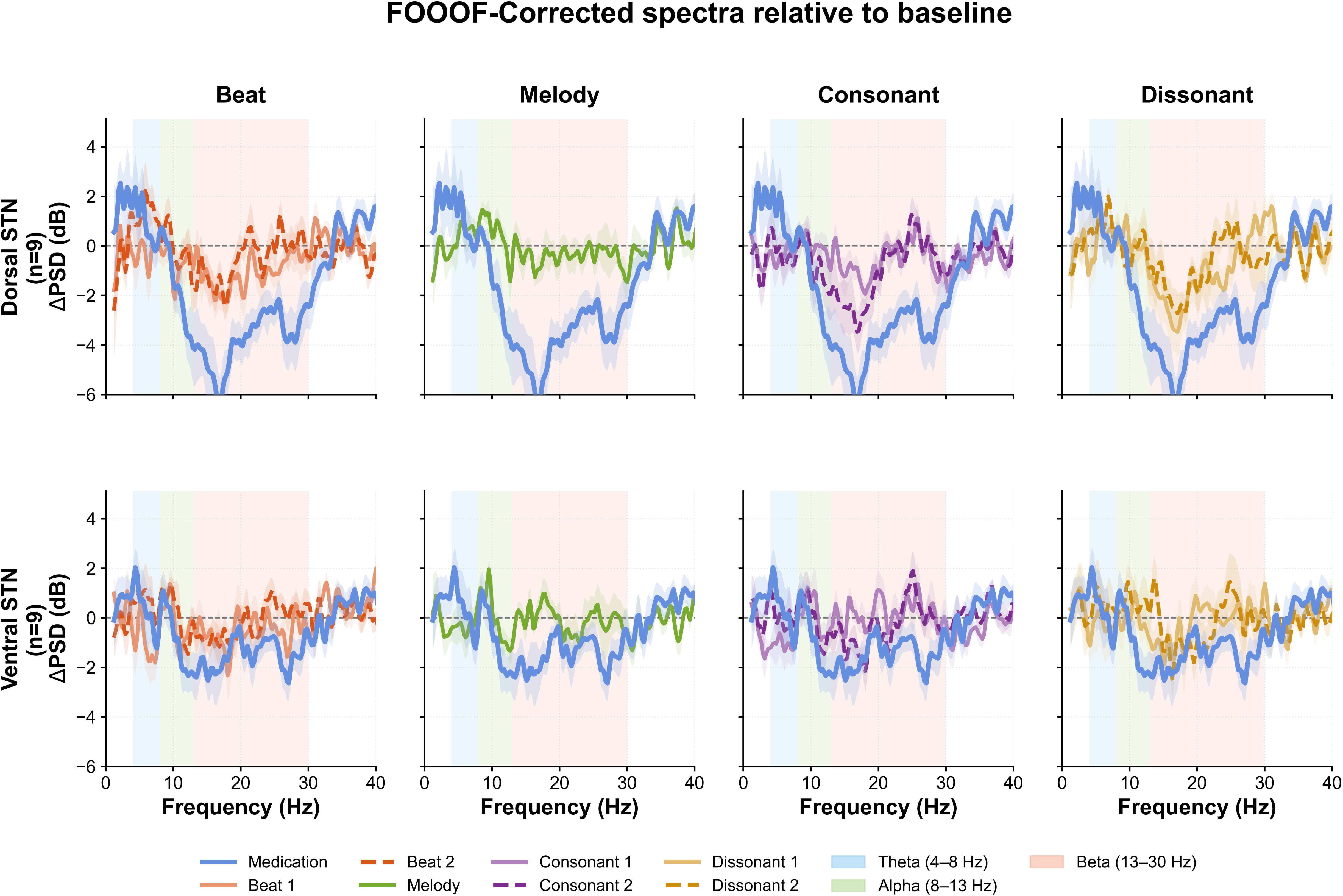
FOOOF-corrected changes in oscillatory spectral power relative to baseline across music conditions and recording sites. Group-averaged aperiodic-corrected power spectral density changes (ΔPSD, dB) are shown for the dorsal (top row) and ventral (bottom row) subthalamic nucleus (STN). Each panel compares the medication- on condition (blue) with one music condition: Beat 1 (60 BPM), Beat 2 (120 BPM), Melody, Consonant 1 (52 BPM), Consonant 2 (120 BPM), Dissonant 1 (52 BPM), or Dissonant 2 (110 BPM). Shaded regions denote the theta (4–8 Hz), alpha (8–13 Hz), and beta (13–30 Hz) frequency bands. Solid lines represent the group mean and shaded envelopes indicate the standard error of the mean (SEM).

Group-averaged FOOOF-corrected ΔPSD showed region- and condition-specific modulation across the seven music conditions (Fig. 2). In dorsal STN, medication-on produced the strongest, most spectrally focal beta suppression, with a trough at 15–18 Hz. Relative to baseline, music beats (60, 120 BPM), consonants (52, 120 BPM), and dissonants (52, 110 BPM) showed the most medication-like profiles, with broad beta reductions; faster beat, consonant, and dissonant stimuli produced more focused lower-beta (13–20 Hz) suppression. Melody caused only modest change, with minimal beta suppression. Lower-tempo beat 1 (60 BPM), consonant 1 (52 BPM), and dissonant 1 (52 BPM) produced intermediate beta reductions, while faster dissonant 2 (110 BPM) caused broader but shallower suppression across 13–30 Hz. Across conditions, activity below ∼8–10 Hz generally showed small positive deviations from baseline, varying by stimulus.

In ventral STN, the spectral profile was similar but much weaker. Beta suppression was reduced across all music conditions and medication-on, with smaller negative deflections than dorsal STN. Inter-patient variability was also greater, reflected by broader SEM envelopes across much of the spectrum. As a result, differences between music conditions were less distinct, though faster-tempo beat and consonant stimuli still most closely resembled the medication-on state compared with melody or slower harmonic conditions.

### FOOOF Band Power Statistics Across Frequency Bands

To assess frequency-specific effects of dopaminergic medication and musical features on subthalamic activity, we measured aperiodic-corrected power in four canonical bands—theta (4–8 Hz), alpha (8–13 Hz), beta (13–30 Hz), and gamma (30–45 Hz); Fig.3. The expanded paradigm dissociated rhythmic (Beat1: 60 BPM; Beat2: 120 BPM), melodic, consonant harmonic (Consonant 1: 52 BPM; Consonant 2: 120 BPM), and dissonant harmonic (Dissonant 1: 52 BPM; Dissonant 2: 110 BPM) components, allowing us to test whether modulation depended on tempo, melody, or harmony.

**Figure 3.**
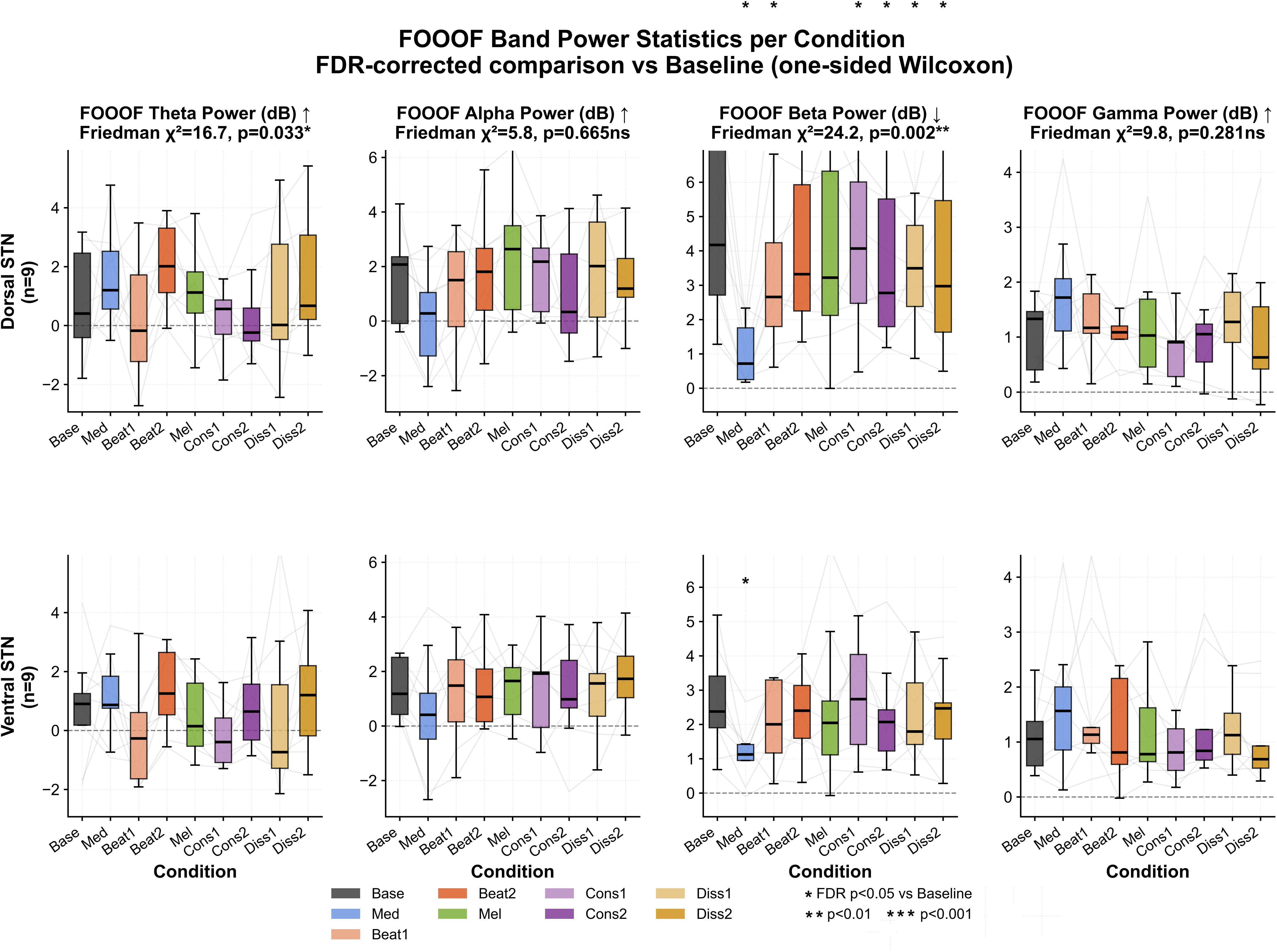
Statistical comparison of the FOOOF corrected mean band powers across all bands for both dorsal and ventral STN recordings.

Effects were evaluated with Friedman tests, followed by FDR-corrected one-sided Wilcoxon tests versus baseline and two-sided paired tests between music and medication.

### Beta band (13–30 Hz): strongest and most consistent modulation

Beta showed the clearest condition-dependent effects, especially in dorsal STN, supporting beta as the main frequency through which structured auditory stimulation interacts with basal ganglia circuits (Fig. 3). In dorsal STN, beta varied significantly across conditions (Friedman χ²(7)=24.18, p=0.0021). Medication strongly suppressed beta relative to baseline (p=0.0156, r=−1.00), and several music conditions also significantly reduced beta after FDR correction: Beat1 (p=0.0156, r=−0.91), Consonant 1 (p=0.0219, r=−0.82), Consonant 2 (p=0.0156, r=−0.96), Dissonant 1 (p=0.0365, r=−0.73), and Dissonant 2 (p=0.0219, r=−0.82). Beat2 and melody showed decreases that did not survive correction. Although significant music effects were large (r=−0.73 to −0.96), paired tests showed medication suppressed beta more than every music condition after FDR correction (Beat1: p=0.0137, r=+0.91; Beat2: p=0.0091, r=+1.00; Consonant1: p=0.0091, r=+1.00; Consonant2: p=0.0137, r=+0.96; Dissonant1: p=0.0091, r=+1.00; Dissonant2: p=0.0137, r=+0.91). Thus, music and medication converge on beta suppression, but music engages overlapping rather than equivalent mechanisms. Because suppression occurred across rhythmic, consonant, and dissonant stimuli, the key driver was unlikely to be tempo or pleasantness alone; instead, structured temporal organization and predictable auditory patterning appear central.

In ventral STN, beta modulation was weaker but still condition-dependent (Friedman χ²(7)=15.91, p=0.0437). Medication reduced beta relative to baseline (p=0.0312, r=−0.96), but no music condition survived baseline correction. Several music conditions nevertheless differed from medication: Beat2 (p=0.0273, r=+1.00), Consonant1 (p=0.0410, r=+0.91), Dissonant1 (p=0.0456, r=+0.87), and Dissonant2 (p=0.0479, r=+0.82). This indicates less consistent music-related beta engagement in ventral than dorsal STN, consistent with stronger dorsal links to sensorimotor timing networks and stronger ventral links to associative/limbic circuits.

### Theta band (4–8 Hz): condition-dependent low-frequency modulation

Theta showed significant overall condition effects in both dorsal and ventral STN. In dorsal STN, theta varied across conditions (Friedman χ²(7)=16.71, p=0.0333), but no individual contrast survived FDR correction. Beat2 and Dissonant2 showed moderate-to-large positive effects (both r=+0.69), suggesting increased theta with faster rhythmic stimulation, whereas consonant conditions showed negative directional effects, implying sensitivity to specific stimulus structure rather than a uniform music effect. Ventral STN showed an even stronger overall theta effect (Friedman χ²(7)=22.16, p=0.0046), again without any individual baseline comparison surviving correction. Together, these findings suggest theta reflects distributed sensitivity to auditory context, possibly involving prediction and timing-related basal ganglia networks.

### Other bands

Alpha (8–13 Hz) and gamma (30–45 Hz) showed no significant condition effects in dorsal STN (alpha: χ²(7)=5.84, p=0.6655; gamma: χ²(7)=9.78, p=0.281) or ventral STN (alpha: χ²(7)=6.28, p=0.6157; gamma: χ²(7)=15.41, p=0.0517). Their relative stability argues against generalized spectral shifts and suggests that effects were confined mainly to bands linked to basal ganglia motor regulation (beta) and temporal coordination (theta).

### Neural state space dynamics: music as a trajectory toward the therapeutic state

The analyses above treated theta and beta as separate measures, but they are not physiologically independent: beta suppression and theta enhancement both reflect reorganization of basal ganglia dynamics under altered dopaminergic tone (Oswal et al., 2013), and their joint pattern may capture effects missed by univariate tests. Clinically, the key question is not only whether music changes individual bands, but whether it shifts the brain toward the oscillatory state produced by dopaminergic medication—the established therapeutic reference in PD. We therefore embedded the same time-resolved band-power estimates into a two-dimensional neural state space defined by joint changes in theta (Δθ) and beta (Δβ) relative to baseline, directly extending the time-frequency analysis into a geometric framework referenced to the pharmacological therapeutic state.

For each patient and condition, each time point was represented as (Δθ, Δβ), with baseline anchored at the origin. Medication defined a reference cluster representing the therapeutic state. The diagonal Δθ = Δβ marks equal co-modulation; the region below it indicates proportionally greater theta enhancement relative to beta suppression, and points above indicate the converse. Euclidean distances between cluster centroids quantified both displacement from baseline and similarity of each music condition to medication, while accounting for covariance structure.

The dorsal STN showed the clearest separation between baseline and stimulation conditions (Fig. 4). Baseline clustered tightly around the origin, whereas all music conditions shifted the neural state toward positive Δθ and negative Δβ values, with medication producing the largest shift (Fig. 4A). Medication was farthest from baseline (d = +1.95, p<0.001), indicating robust reorganization under dopaminergic treatment. Among music conditions, Beat1 (d = +1.09), Beat2 (d = +1.00), Consonance2 (d = +0.91), and Dissonance2 (d = +0.99) produced the largest displacements; Melody was moderate (d = +0.63), Consonance1 was small (d = +0.26), and Dissonance1 was intermediate (d = +0.70). Thus, rhythmic stimulation and stronger harmonic conditions most closely approached the medication-induced state. The zoomed version of the Fig. 4A is as shown in Fig. 4B.

**Figure 4.**
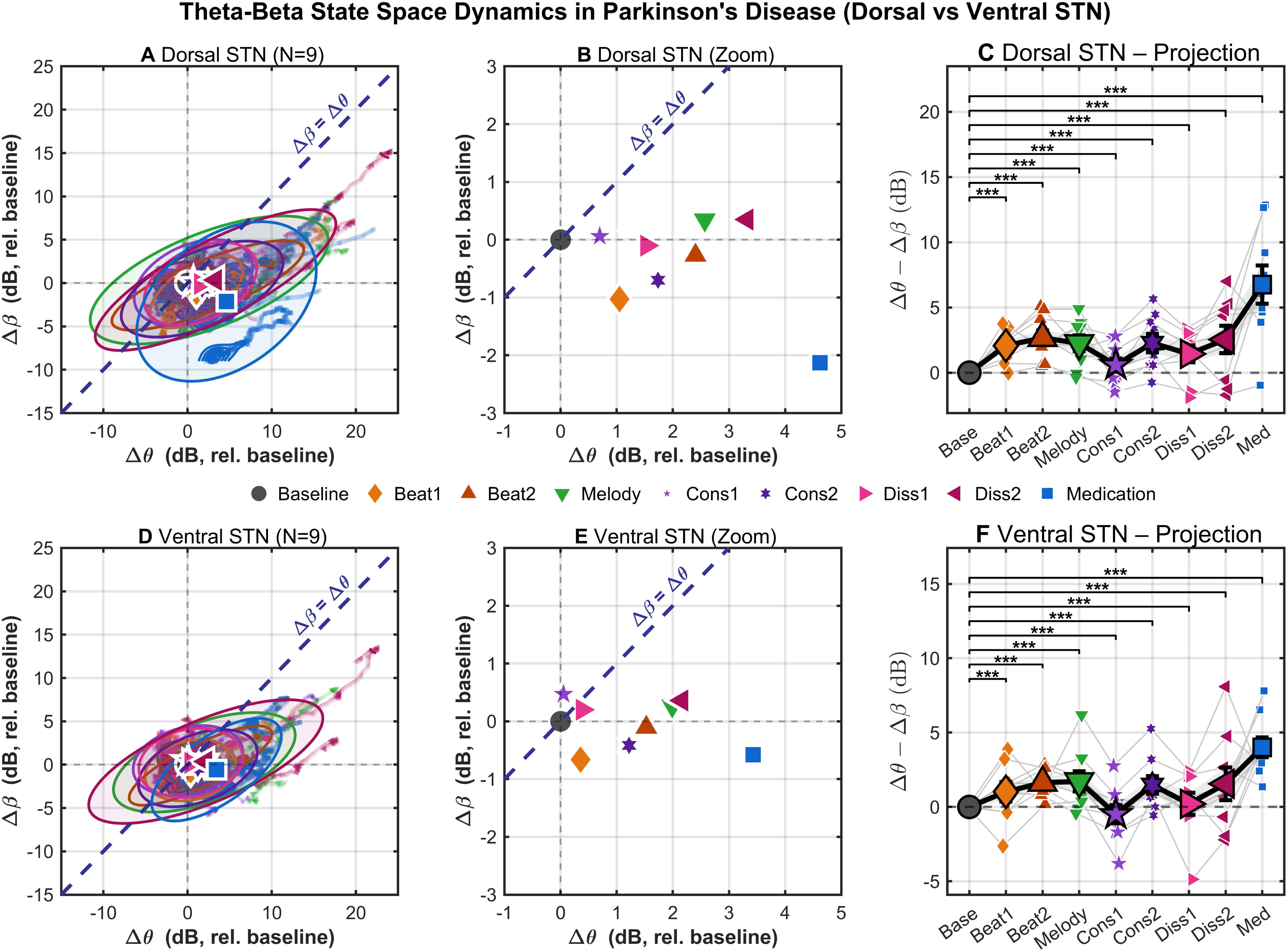
Neural state-space dynamics of theta-beta oscillatory activity in dorsal and ventral STN. (A,D) Two-dimensional neural state space defined by changes in theta (Δθ, x-axis) and beta (Δβ, y-axis) power relative to the OFF-medication baseline for dorsal (A) and ventral (D) STN contacts (N = 9). Colored trajectories represent the evolution of each musical condition and medication, while ellipses denote the covariance of the corresponding state distributions. The dashed diagonal indicates the line of equal theta and beta modulation (Δθ = Δβ). Insets summarize standardized effect sizes (Cohen’s d) relative to baseline for each condition. (C,F) Projection of each condition onto the Δθ − Δβ axis, summarizing the balance between theta and beta modulation. Individual participant values are shown in color, with black symbols representing group means ± SEM. Medication produced the largest positive projection in both dorsal and ventral STN, whereas rhythmic and selected harmonic conditions generated intermediate shifts toward the medication-associated neural state. Statistical significance is indicated by *** (p < 0.001). (B,E) are the zoom in versions of (A,D) respectively. Modulatory effect at dorsal STN: Medication (d = +1.95, p<0.001), Beat1 (d = +1.09), Beat2 (d = +1.00), Consonance2 (d = +0.91), and Dissonance2 (d = +0.99), Melody (d = +0.63), Consonance1 (d = +0.26), and Dissonance1 (d = +0.70). Modulatory effect at ventral STN: Medication again produced the largest displacement from baseline (d = +1.79, p<0.001), Beat2 (d = +0.76), Consonance2 (d = +0.68), Melody (d = +0.58), Beat1 (d = +0.46) and Dissonance2 (d = +0.54), Dissonance1 (d = +0.08), and Consonance1 (d = −0.17).

Projection analysis (Fig. 4C) clarified this further. Medication produced the largest positive Δθ − Δβ projection, significantly exceeding every music condition (all significant, p<0.001). Still, Beat2, Melody, Consonance2, and Dissonance2 consistently produced positive projections across patients, indicating coordinated theta-dominant modulation in the same direction as medication. Beat1 showed a smaller but still positive shift, whereas Consonance1 remained near baseline, suggesting weak engagement of the therapeutic oscillatory axis. Although individual trajectories varied, the ordering of conditions was highly consistent, with medication producing the largest and most reliable movement in state space.

A similar but weaker pattern appeared in ventral STN (Fig. 4D). Medication again produced the largest displacement from baseline (d = +1.79, p<0.001), showing that dopaminergic therapy reorganizes oscillatory dynamics throughout the STN. Music- induced shifts were smaller than dorsally and remained closer to baseline. Beat2 (d = +0.76), Consonance2 (d = +0.68), and Melody (d = +0.58) produced the strongest ventral responses; Beat1 (d = +0.46) and Dissonance2 (d = +0.54) were moderate. Dissonance1 showed minimal displacement (d = +0.08), and Consonance1 was the only condition with a slight negative effect (d = −0.17), indicating little or no movement away from baseline. The zoomed in version of the Fig. 4D is shown in Fig. 4E. Projection analysis (Fig. 4F) likewise showed that medication produced the greatest positive Δθ − Δβ value. Most music conditions produced modest positive projections, with Beat2, Melody, and Consonance2 showing the clearest shift toward the medication-associated axis. However, inter-patient spread was larger than in dorsal STN, and several patients showed little movement during music despite robust medication responses, consistent with weaker overall ventral effect sizes.

Overall, these findings show that dorsal STN is robustly responsive to music, with rhythmic structure and stronger harmonic manipulations producing the strong convergence toward the pharmacological neural state. Ventral STN also participates in music-related network reorganization, but its responses are much smaller and less consistent than those in the dorsal sensorimotor territory.

### Cortical drive to basal ganglia during music: Phase Slope Index

The preceding analyses established that music induces frequency-specific and region-dependent reorganizations of STN oscillatory power, with beta suppression and theta enhancement representing the primary and secondary effects respectively. However, these power analyses do not address the mechanism by which an auditory stimulus modifies subcortical LFP dynamics. A key mechanistic question is whether the observed STN changes are driven by top-down cortical input consistent with the cortico-subthalamic hyperdirect pathway (Nambu et al., 2002) or reflect local subcortical reorganization. To address this, we computed the Phase Slope Index (PSI; Nolte et al., 2008) between scalp EEG and STN LFP signals across all frequency bands, quantifying the directionality of information flow between cortex and STN (Fig. 5). The music-induced change in directionality (ΔPSI = PSI_music − PSI_baseline) was used to identify connectivity shifts specifically attributable to music exposure.

**Figure 5.**
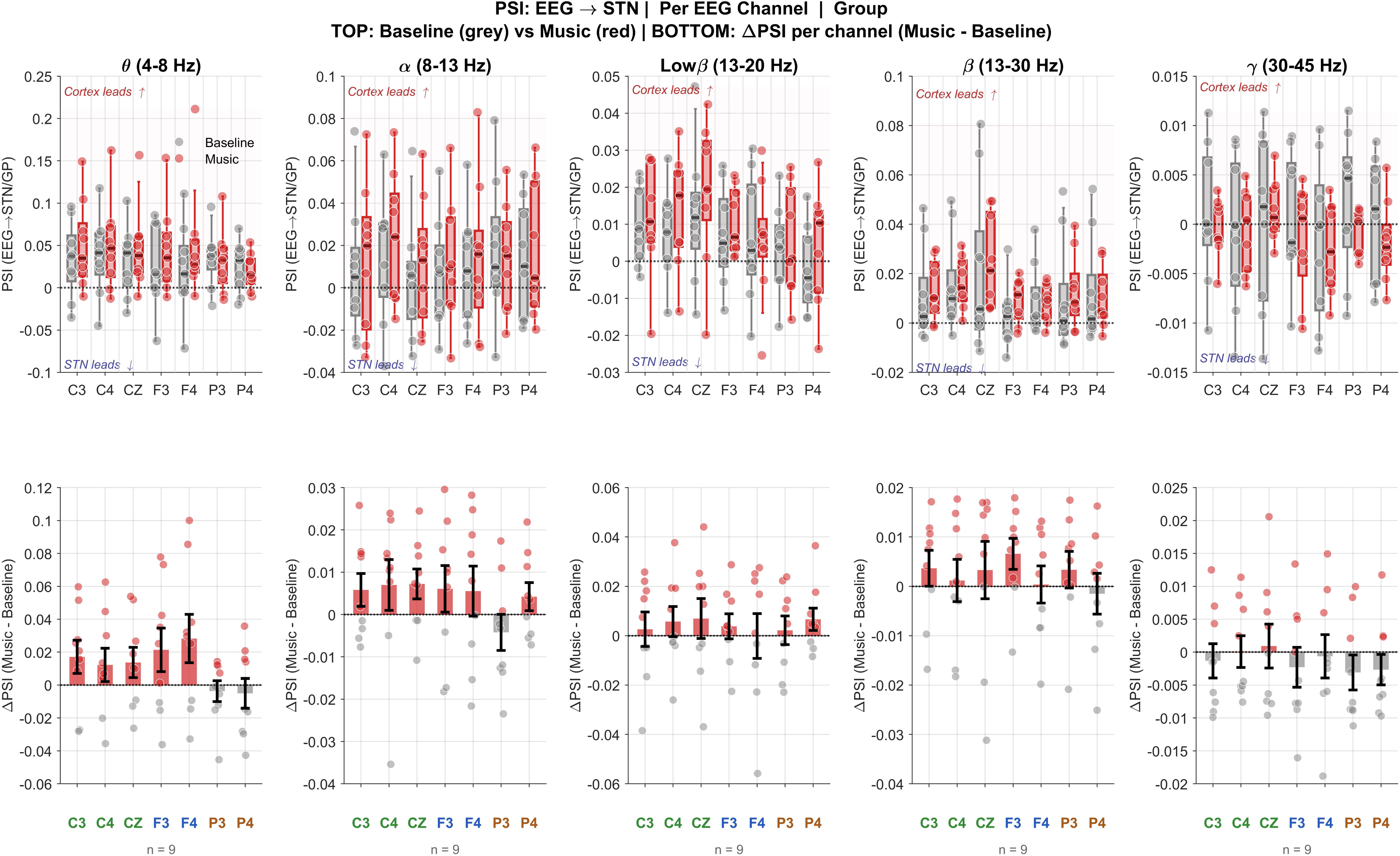
Dorsal STN: Cortical–subcortical interactions reveal cortex-driven reorganization of basal ganglia dynamics during music. A) Top panel : Phase Slope Index (PSI) between cortical EEG electrodes and subthalamic nucleus (STN) LFPs, shown as music-induced change in directionality (Δ*PSI* = *PSI_Music_* − *PSI_Baseline_*). across frequency bands and recording sites. Positive values indicate cortex → STN influence; negative values indicate STN → cortex influence. B) Bottom panel: Positive ΔPSI values (warm colors) indicate cortex → STN drive, whereas negative values (cool colors) indicate STN → cortex influence.

### Low Beta Band (13–20 Hz): Music Enhances Cortical Drive to STN

The most robust and consistent was a music-induced increase in cortex-to-STN directional connectivity in the low beta band (13–20 Hz). In dorsal STN (Fig. 6), ΔPSI was showing strong positive at C4 during music in the low beta band region along with a strong positive trend at F4 in the theta band, indicating that cortex increased their directional drive to STN specifically during music exposure relative to spontaneous baseline. This enhancement of cortical drive occurred precisely in the frequency band and the STN subregions showing the strongest and most consistent beta suppression in Fig. 2, 3 along with associated theta enhancement. The temporal coincidence of increased cortex→STN directionality with reduced STN beta power is mechanistically coherent: strengthened top-down cortical input in the beta range may disrupt pathological STN beta synchrony through phase desynchronization, consistent with established models of cortico-subthalamic inhibitory control (Nambu et al., 2002; Kühn et al., 2006).

**Figure 6.**
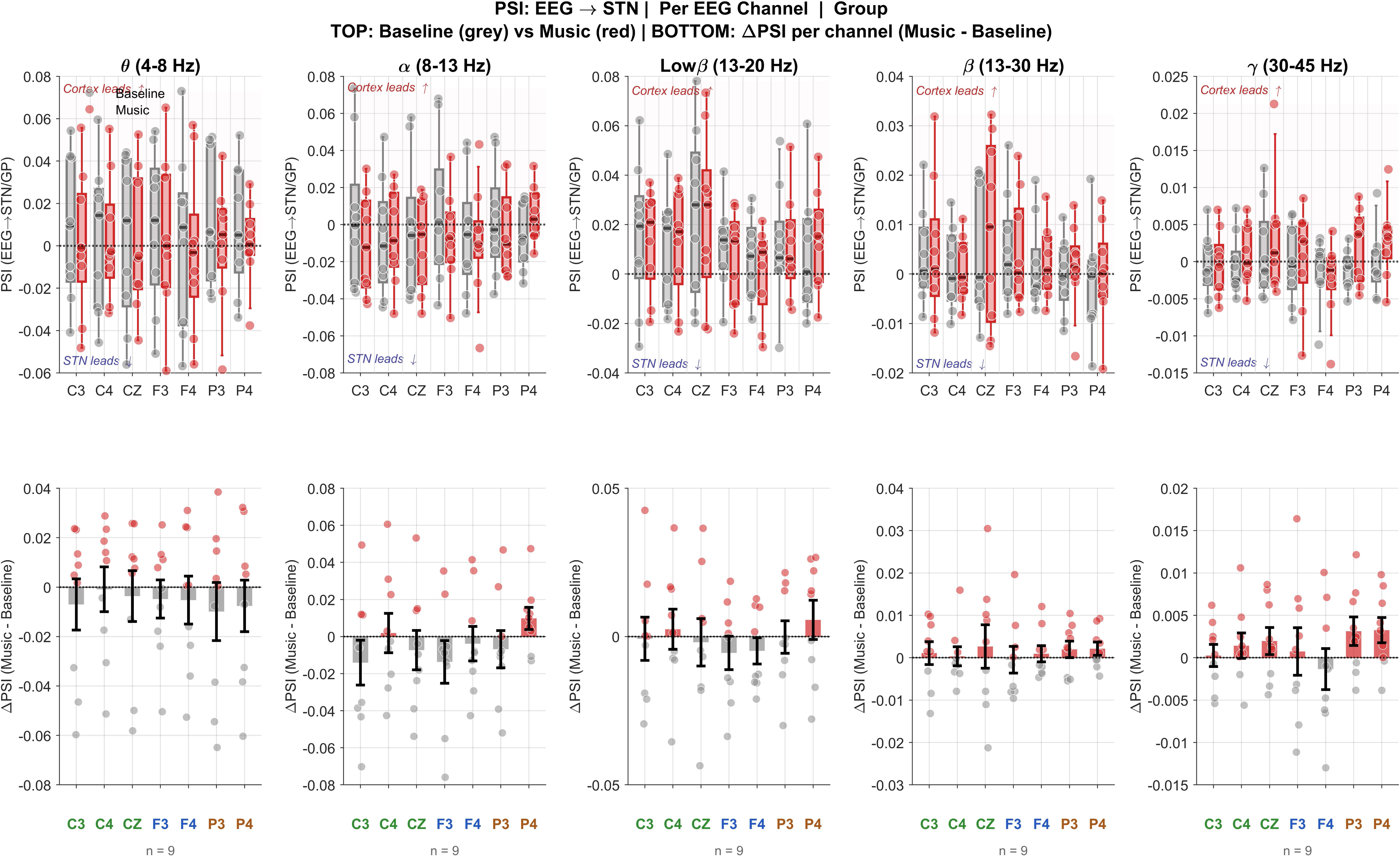
Ventral STN: Cortical–subcortical interactions reveal cortex-driven reorganization of basal ganglia dynamics during music. A) Top panel : Phase Slope Index (PSI) between cortical EEG electrodes and subthalamic nucleus (STN) LFPs, shown as music-induced change in directionality (Δ*PSI*). across frequency bands and recording sites. Positive values indicate cortex → STN influence; negative values indicate STN → cortex influence. B) Bottom panel: Positive ΔPSI values (warm colors) indicate cortex → STN drive, whereas negative values (cool colors) indicate STN → cortex influence.

In ventral STN (Fig. 6), the low beta ΔPSI pattern was directionally consistent with dorsal STN, with positive values at C3 and C4, with attenuated but similar effects. The convergence of this finding across both subregions strengthens confidence in its reliability, despite the weaker univariate power effects observed in ventral contacts.

A distinct finding emerged at parietal channels (P3, P4): ΔPSI in the low beta band was negative at these electrodes across both STN subregions, indicating that during music, STN activity tended to lead parietal cortex rather than the reverse. This contrasts with the positive cortex-leads-STN directionality at frontal and central channels and may reflect feedback signaling through thalamo-cortical loops projecting to posterior parietal areas involved in sensorimotor integration (Haynes & Haber, 2013). This bidirectional reconfiguration - cortex driving STN at frontal/motor sites, STN driving parietal cortex suggests that music-induced network reorganization involves simultaneous top-down and feedback signaling rather than a simple unidirectional cascade.

### Theta band (4–8 Hz): region-specific cortical drive mirrors power effects

In the theta band, a music-induced positive ΔPSI at frontal and central channels was present in dorsal STN (Fig. 6), indicating that cortex tended to lead STN in theta during music at F3, F4, C3, and C4. This is consistent with the dorsal STN theta power trend (Fig. 3) and suggests that the theta effect may be cortically driven: frontal theta generated during auditory attention and predictive processing of musical rhythm (Cavanagh & Frank, 2014) could propagate to STN via the hyperdirect pathway (Nambu et al., 2002), entraining local theta oscillations.

Critically, this theta directionality pattern was absent in ventral STN (Fig. 6), where ΔPSI in the theta band was near zero or negative across all channels. This is precisely consistent with the absence of significant theta enhancement in ventral contacts in Fig. 3 and provides a mechanistic explanation at the connectivity level: without enhanced cortical theta drive, ventral STN contacts do not show reliable theta modulation during music. This spatial dissociation of theta connectivity present in dorsal, absent in ventral mechanistically reinforces the spatial dissociation of theta power effects established in the univariate analyses.

## Discussion

An important open question in PD research is whether non-pharmacological interventions can produce neural effects that meaningfully approximate dopaminergic therapy. Music has emerged as a promising adjunct intervention in PD rehabilitation, yet its neural equivalence to pharmacological treatment and the mechanisms by which it engages disease-relevant circuitry have remained unclear. Here, using postoperative LFP recordings from the STN, we directly compared music and medication-induced modulation of subcortical oscillatory activity across seven systematically designed musical conditions that independently varied tempo, melodic structure, and harmonic valence. The results demonstrate that structured musical stimulation can partially but measurably recapitulate the neural signatures of dopaminergic therapy, and that tempo rather than harmonic pleasantness — is the primary acoustic determinant of basal ganglia engagement.

### The Pattern of Beta Suppression as a Shared Mechanism

Across all analyses, beta suppression emerged as the dominant and statistically robust finding, present across five of seven music conditions in dorsal STN and under medication in both STN subregions. The highly significant Friedman omnibus effect in dorsal STN confirms that condition-dependent beta modulation was systematic and reproducible across patients rather than driven by individual variability. This establishes beta modulation as the primary shared oscillatory mechanism linking structured auditory stimulation and dopaminergic therapy, consistent with the established role of beta synchrony as the canonical biomarker of the hypodopaminergic state in PD (Hammond et al., 2007; Little & Brown, 2014).

However, the magnitude of beta modulation clearly distinguished music from medication. Dopaminergic treatment consistently produced greater beta suppression than any individual musical condition, indicating that music engages a related but weaker mechanism rather than reproducing the full electrophysiological effect of pharmacological restoration. The state-space analysis further demonstrated this graded relationship, with the most effective musical conditions approaching but not reaching the medication-associated oscillatory state. Together, these findings support the interpretation that music produces meaningful convergence with dopaminergic neural dynamics while remaining an adjunctive rather than equivalent intervention.

### Tempo, Not Harmonic Valence, as the Organizing Principle

These findings are consistent with models of basal ganglia entrainment, in which rhythmic auditory input can synchronize motor cortico-subcortical networks when temporal regularity aligns with the dynamics of movement-related circuits (Large & Snyder, 2009; Grahn & Brett, 2007). Faster tempos in the 100–120 BPM range overlap with typical human walking cadence and have previously been associated with effective motor entrainment during rhythmic auditory stimulation (Thaut et al., 1996; Thaut & Hoemberg, 2015). Our findings extend these observations by showing that similar temporal features are associated with greater convergence of STN oscillatory dynamics toward the medication-associated state.

### Harmonic Valence Is Not a Barrier: The Dissonance Finding

A second theoretically important finding is that dissonant harmonic conditions (Dissonant1 and Dissonant2) produced significant beta suppression in dorsal STN — Dissonant1 and Dissonant2 comparable in magnitude to consonant conditions at matched tempo. This directly demonstrates that harmonic pleasantness or affective valence does not gate the beta suppression effect. Dissonance and consonance, which differ in their recruitment of limbic and reward circuitry (Blood & Zatorre, 2001; Salimpoor et al., 2011), produce indistinguishable oscillatory consequences in the STN when tempo is matched. This dissociation between subjective musical valence and subcortical motor-relevant oscillatory change may implicate music therapy design: it suggests that the neural mechanisms driving beta suppression operate on the temporal structure of the auditory input rather than on its affective or tonal quality. Patients with PD who dislike or are unfamiliar with harmonically consonant music may respond equally well to dissonant stimuli of equivalent tempo, a hypothesis testable in future work.

The likely mechanism is that both consonant and dissonant stimuli at sufficient tempo generate rhythmically structured cortical activity that propagates to STN via the hyperdirect cortico-subthalamic pathway (Nambu et al., 2002), regardless of the harmonic content carried by that rhythm. The auditory-motor coupling that drives this entrainment depends primarily on the periodicity and predictability of the temporal envelope, which is shared between consonant and dissonant versions at matched beats per minute.

### Melody: Moderate and Tempo-Independent

The melody produced intermediate effects at the state-space level (dorsal d = 0.63; ventral d = 0.58), ranking above the slow harmonic conditions but below all fast-tempo rhythmic and harmonic conditions. Importantly, the melody did not contain an explicit tempo manipulation and is therefore not directly comparable to the tempo-contrasted pairs. Its intermediate position likely reflects the engagement of auditory-motor integration networks through pitch contour processing and melodic prediction (Zatorre et al., 2002), which may indirectly recruit motor-relevant cortical areas without the temporal specificity of a periodic beat. The absence of a statistically significant beta suppression effect after FDR correction in dorsal STN, despite a moderate state-space displacement, suggests that melodic processing provides distributed but less targeted engagement of basal ganglia beta regulation networks than structured rhythmic stimulation.

### Theta Enhancement: Condition-Dependent and Broadly Distributed

Theta oscillations showed significant omnibus condition effects in both dorsal and ventral STN, indicating that low-frequency dynamics were sensitive to the expanded musical stimulus space. However, no individual condition survived FDR correction in either region. In dorsal STN, directional theta enhancement was most prominent under Beat2 and Dissonant2 (both r = +0.69), again implicating higher-tempo stimulation.

Notably, consonant conditions showed negative or near-zero directional effects on theta, suggesting that harmonic richness without strong rhythmic drive does not consistently enhance theta activity. This is consistent with the hypothesis that theta modulation is driven by temporal prediction circuits rather than by tonal or affective processing per se (Cavanagh & Frank, 2014; Grahn & Rowe, 2009).

The significant ventral STN Friedman effect for theta despite no individual condition surviving correction indicates distributed sensitivity to the auditory context rather than a selective, condition-locked response. This may reflect the engagement of associative and limbic STN territories in broader auditory processing and attentional orienting, rather than motor-specific theta entrainment.

Theta oscillations showed significant overall condition effects in both dorsal and ventral STN, indicating that low-frequency dynamics were sensitive to the broader musical stimulus space. However, no individual condition survived FDR correction, suggesting that theta modulation was weaker and less stimulus-specific than the beta effects. In dorsal STN, the largest directional theta increases occurred during Beat2 and Dissonant2, suggesting a possible sensitivity to faster rhythmic stimulation. Conversely, consonant conditions showed minimal or negative directional changes, raising the possibility that harmonic structure alone is insufficient to consistently enhance theta activity. This pattern is consistent with a potential contribution of temporal prediction and timing-related networks to theta modulation, rather than a primary dependence on tonal or affective features (Cavanagh & Frank, 2014; Grahn & Rowe, 2009).

In ventral STN, the significant omnibus theta effect in the absence of significant individual contrasts suggests broader contextual sensitivity rather than a selective response to specific musical features. This may reflect engagement of associative and integrative STN networks involved in processing auditory context and attention, rather than the more sensorimotor-specific modulation observed in dorsal STN.

### Alpha and Gamma: Frequency Selectivity of Music-Induced Modulation

Alpha and gamma power remained stable across all conditions and regions confirming that the observed beta and theta effects represent frequency-selective modulation rather than a generalized spectral shift. The absence of consistent gamma enhancement with music, in contrast to the positive gamma trend observed under medication in prior analyses, suggests that music does not recapitulate the dopamine-dependent fast interneuron modulation that drives pharmacological gamma enhancement (Brücke et al., 2008). This dissociation also demonstrates that music is a frequency-selective, not global, pharmacological mimic: convergence with medication occurs primarily in the beta band and to a lesser extent in theta, while the fast oscillatory range diverges.

### The State Space as a Unified Quantitative Framework

The clear ordering of conditions across both STN subregions — medication > fast-tempo rhythmic/harmonic conditions > melody > slow harmonic conditions — demonstrates that state-space displacement is graded and condition-specific, not a binary music/no-music distinction. The metric of music-to-medication displacement ratio (50–56% for the most effective conditions) provides a quantifiable endpoint for comparing music therapy protocols and could in principle serve as an objective neural benchmark for intervention optimization in future work. The state-space analysis transforms the evaluative question from “does music change the brain?∝ to “does music move the brain toward the state associated with clinical benefit?∝ The answer is consistently affirmative across both STN subregions. All music sub-components displaced neural activity in the therapeutically relevant direction, with beat and consonant stimulation achieving the greatest proximity to the medication cluster. The incomplete overlap between music and medication states, and the significant pairwise distances that persist even for the most effective music sub-components, is consistent with the univariate finding that medication produces larger effect sizes across all bands. This partial convergence indicates that music engages similar circuit mechanisms to dopaminergic therapy but does so with reduced magnitude and through partially distinct spectral pathways.

### Cortical Mechanism: Top-Down Drive via the Hyperdirect Pathway

The PSI analysis supports a mechanistic account in which structured musical stimulation engages auditory-motor cortical networks that generate rhythmically organized activity in the low beta and theta frequency ranges. This cortical activity propagates to STN via the hyper direct cortico-subthalamic pathway (Nambu et al., 2002; Haynes & Haber, 2013), providing rapid top-down influence over pathological basal ganglia synchrony. The significant positive ΔPSI at contralateral motor (C4) and frontal (F4) channels during music in the low beta band — the same band showing the strongest STN power suppression — is consistent with enhanced cortical drive disrupting pathological beta synchrony through phase desynchronization, an established mechanism in cortico-subthalamic inhibitory control (Kühn et al., 2006). The absence of theta directionality in ventral STN, mirroring the absence of significant theta power modulation in that region, further supports the interpretation that both beta and theta effects are cortically driven through topographically specific projections rather than arising from local subcortical reorganization (Haynes & Haber, 2013).

The STN-leads-parietal directionality (negative ΔPSI at P3, P4) observed across both subregions suggests that music-induced network reorganization is not a simple unidirectional cortex-to-STN cascade but involves simultaneous feedback signaling through thalamo-cortical loops to posterior parietal areas. This bidirectional reconfiguration may represent a broader sensorimotor integration process recruited during rhythmic auditory processing (Zatorre et al., 2007; Chen et al., 2008).

Given the tempo-dependence of the power and state-space findings, an important prediction follows for the PSI mechanism: higher-tempo conditions should show stronger and more channel-specific ΔPSI than lower-tempo conditions. Testing this prediction in future studies with per-condition PSI analysis would provide direct connectivity evidence for the tempo-entrainment hypothesis and identify the specific cortical channels most effective in driving subcortical state transitions. Auditory-motor cortical networks are known to be engaged during music perception even during passive listening (Zatorre et al., 2007; Chen et al., 2008), and rhythmic auditory stimulation has been shown to modulate basal ganglia-cortical networks implicated in movement and timing (Grahn & Brett, 2007; Thaut & Hoemberg, 2015).

### A Multi-Route Model Centered on Temporal Structure

Taken together, the seven-condition paradigm supports a model in which temporal structure appears to be the dominant organizing feature of music-induced basal ganglia modulation. Faster, rhythmically regular stimuli were associated with greater movement toward the medication-associated oscillatory state, suggesting that predictive temporal entrainment may provide a particularly effective route for engaging motor cortico-subthalamic networks. Importantly, this effect was observed for both consonant and dissonant stimuli within the tested paradigm, indicating that harmonic valence does not appear to be a necessary condition for STN engagement.

Melodic content produced intermediate effects, likely through auditory-motor integration and predictive processing, but did not achieve the same degree of beta modulation observed with stronger rhythmic structure. Conversely, slower harmonic stimuli provided weaker temporal organization and produced the smallest neural-state shifts. Together, these findings suggest a multi-route model in which different musical features engage basal ganglia circuits through partially overlapping pathways, with temporal regularity providing the strongest influence on STN beta dynamics.

This framework has implications for future music-based interventions in PD. Rather than if harmonic preference alone determines neural efficacy, optimization strategies may benefit from prioritizing temporal structure and rhythmic predictability while allowing flexibility in musical content. Future studies should directly test whether tempo systematically influences beta modulation and whether these neural effects translate into measurable behavioral and clinical benefits.

### Clinical Implications, Limitations to be Addressed in Future Studies

The present findings provide electrophysiological evidence that structured musical stimulation engages basal ganglia circuits also influenced by pharmacological therapy in PD, with temporal structure emerging as a key feature associated with STN modulation. Across multiple musical conditions, basal ganglia dynamics shifted toward the medication-associated oscillatory state, and increased cortical influence on STN provided a plausible mechanistic framework for these effects. The comparable neural effects of consonant and dissonant stimuli at matched tempos suggest that effective musical stimulation may not require a specific harmonic valence, potentially allowing greater flexibility in individualized music-based interventions. Future work should determine whether the degree of neural-state convergence achieved by music is sufficient to produce measurable motor benefits in the off-medication state and whether STN responses scale systematically with tempo.

Several limitations constrain interpretation but also define priorities for future studies. The sample size was small; recordings were obtained acutely after surgery with externalized electrodes; stimuli were standardized rather than individualized according to preference or familiarity; no concurrent behavioral measures were collected; PSI reflects directional functional interactions rather than anatomical connectivity; and connectivity analyses were not resolved by individual stimulus condition. These limitations mean that the present findings should be considered a proof-of-concept, while providing a foundation for larger studies incorporating behavioral outcomes, personalized stimuli, and condition-specific connectivity analyses.

## Data Availability

All MATLAB and Python scripts used for analyzing the data is uploaded in the GitHub. This can be provided on reasonable requests.

## Data Availability

All data produced in the present study are available upon reasonable request to the authors.

## Acknowledgements

Authors thank Prajakta Joshi, PhD for technical support.

